# A Hybrid Framework for Accurate Melanoma Diagnosis: Leveraging Generative AI with Enhanced CNN+ Architectures

**DOI:** 10.64898/2026.04.27.26351813

**Authors:** Yiming Wu, Bojun Zhang, Yan Yan, Jiashu Li, Yue Wu, Sung Soo Kim, Kuan Huang, Qiong Ye, Yingzhe Yu, Guanchao Tong

**Affiliations:** College of Science, Mathematics and Technology, Wenzhou-Kean University, 88 Daxue Rd, Ouhai, Wenzhou, Zhejiang Province 325060, China; College of Science, Mathematics and Technology, Kean University, 1000 Morris Avenue, Union, NJ 07083, USA; Department of Dermatology, The First Affiliated Hospital of Ningbo University, No. 59 Liuting Street, Ningbo, Zhejiang Province 315010, China; GCTB–NSU Joint Institute of Technology, Guangzhou College of Technology and Business, Foshan, Guangdong Province 528138, China

**Keywords:** deep/machine learning for feature extraction, image classification models

## Abstract

Melanocytes become cancerous, forming tumors that may invade and destroy the surrounding tissues. When melanocytes acquire invasive characteristics, the anchored melanoma begins to damage the normal cells. Therefore, early intervention and diagnosis are essential to avoid high morbidity and mortality in malignant melanoma. However, It is challenging to distinguish the difference between malignant melanoma and benign clump of melanocytes. Based on a data set of 10,000 melanocyte tumors, this paper develops a new model system to improve the accuracy of distinguishing between benign and malignant melanocytes. In the first stage, the original CNN architectures are used, such as ResNet18, ResNet50, VGG11, and VGG16. Synthetic medical images, generated via a Diffusion Model to extract informative features from the original dataset, are used to train the CNN architectures. This approach improves classification accuracy from 91.1% to 92.9%. In the second stage, the fully connected layer of each neural network is replaced with a high-level classifier, XGBoost, to perform secondary classification. This hybrid strategy further enhances performance, achieving up to 93.3% accuracy by using the synthetic images.

## I. Introduction

Melanoma is a highly malignant cancer among skin diseases. Since melanin is present in skin cells, abnormal cells containing melanin may migrate or infiltrate different parts of the body [1]. If it spreads to critical organs, it can cause organ failure and even endanger life. Studies have shown that early detection of melanoma can dramatically improve survival and cure rates [2] [3]. In addition, skin cancer can be roughly divided into three types: basal cell skin cancer (BCC), squamous cell skin cancer (SCC), and melanoma. Among them, melanoma has the highest mortality rate. Therefore, it is very important to diagnose these melanoma malignancies at an early stage.

The diagnosis of melanoma depended on the clinical manifestation, dermoscopy images and histopathological confirmation. The diagnostic evaluation in clinical settings is primarily based on the ABCDE criteria: asymmetry (A), border irregularity (B), color variation (C), diameter (D) and evolving size and elevation (E) [4], However, visual inspection of skin lesions largely depends on the physician’s experience and technical proficiency, while the evaluation of dermoscopy images may also be affected by image quality issues. Therefore, excisional biopsy to obtain tissue samples and perform tissue analysis is the benchmark for confirming melanoma. However, excision will leave scars on the skin. In addition, for patients with dysplastic nevus syndrome, it is not realistic to perform whole-body excision for patients with many abnormal cells [5].

In recent years, the rapid rise of AI technology significantly enhanced various fields to many fields, such as transportation, climate change, and biomedicine. In the domain of skin disease diagnosis, AI typically employs two main approaches. The first is the expert system, which formalizes the expert’s knowledge through an understandable framework [6]. This method is highly understandable, but it requires extensive preparation and is labor-intensive. The second usual approach is by utilizing CNN architectures [7] [8], which are trained on labeled melanoma image datasets. However, the limited availability of labeled medical data presents a major challenge. Data scarcity often leads to issues such as overfitting, poor generalization, and weak transferability, all of which hinder the advancement of AI-driven medical diagnosis [9]. Addressing the shortage of melanoma datasets is therefore essential to improving diagnostic accuracy. Additionally, optimizing the neural network architecture can further enhance classification performance [10].

To address these problems, we used a generative model-diffusion model to generate the synthetic image dataset. By using the Diffusion model [11], we generated 8 sets of different size of the images. In addition, we modified the architecture of the traditional neural network model, where we replaced the fully connected layer by another classifier XGBoost. We compared the performance across the different models and found that using the synthetic dataset along with integrating the XGBoost classifier significantly improved the classification accuracy.

## II. Dataset and Methods

### A. Dataset and Image preprocessing

In this study, we utilized a melanoma dataset from Kaggle [12], originally containing images sized at 300×300 pixels, comprising 9,600 training images and 1,000 test images. We resized the images to 64×64 pixels, aligning with the requirements of the Denoising Diffusion Probabilistic Models (DDPM). Despite the reduced resolution, our experiments demonstrated that these smaller images retained robust performance in classification tasks.

### B. Development of the Generative Diffusion Dataset (GDD)

The development of GDD is inspired by generative models, which are frequently employed to address the challenge of insufficient medical imaging data [13] [14]. Using Denoising Diffusion Probabilistic Models (DDPM), we generated synthetic images to extract meaningful information from the original dataset. In our study, DDPM was specifically utilized for data augmentation during dataset construction. We introduced a parameter lambda (*λ*) to represent the relative size of each Generative Diffusion Dataset (GDD), defined as the ratio of synthetic images in the Generative Diffusion Dataset (GDD) dataset to the original training images. When the value of lambda (*λ*) is equal to 0, it means there is not synthetic images in that GDD.

### C. Stage 1 Developing the DDPM-CNN Models

CNN architectures have proven to be very effective in classification tasks [15]. After employing the Denoising Diffusion Probabilistic Model (DDPM) to augment our dataset, it was essential to apply robust deep learning models for melanoma identification. Specifically, we evaluated four widely used CNN architectures for the classification task: ResNet18, ResNet50, VGG11, and VGG16.

The characteristics of these four network architectures are as follows:

VGG11: This network has 11 layers, including eight 3×3 convolutional layers and three fully connected layers. It inherits the simple structure of “unified convolution + pooling” of the VGG series and deepens the network depth without significantly increasing the number of parameters by stacking small convolution kernels, thereby improving the feature expression ability. VGG11 has been used for the task of screening cervical cancer [16].

VGG16: This network contains 41 layers, 16 of which are learnable weights [17]. It has a Top-5 accuracy of 92.77% on the same ImageNet dataset. The input size is the same as 224×224×3 [18]. VGG16 has been shown to have high accuracy in thymic carcinoma diagnosis tasks [19].

ResNet18: This is an 18-layer convolutional neural network that directly transfers input across layers through residual (skip) connections, significantly alleviating the gradient vanishing problem of deep networks and making the network easier to train when the number of layers is increased. ResNet18 has a significant effect in the task of diagnosing rectal cancer [20].

ResNet50: This network consists of 50 layers and uses a residual connection structure to ensure the trainability and convergence speed of deeper models. This residual network structure introduces skip connections and residual mapping to enable more efficient optimization of deep-stacked neural networks, thereby reducing training errors [18]. ResNet50 has been shown to be a model with a higher accuracy rate in pneumonia classification tasks [21].

The Denoising Diffusion Probabilistic Model (DDPM) and the four selected CNN architectures—ResNet18, ResNet50, VGG11, and VGG16—were implemented using PyTorch. Eight Generative Diffusion Datasets (GDDs) are generated by DDPM. The GDDs are used as input to train the ResNet18, ResNet50, VGG11, and VGG16 models. Each model is trained for 100 epochs.

### D. Stage 2 Developing the DDPM-CNN-XGBoost Models

In stage 2, this study removed the outermost fully connected layer in each classic CNN algorithm model and replaced it with the XGBoost classifier. The last fully connected layer was removed in the experiment. When training the XGBoost classifier, we have saved the models of four algorithms during stage 1 training. In stage 2, we directly use the pre-trained model to initialize CNNs with the saved parameters. In the initialization part, we fine-tuned all four algorithm models of stage 1 and obtained the best hyperparameters, which were applied to stage 2 training. Fine-tuning pre-trained convolutional neural network models (CNN) is better than training models from scratch [22]. Following initialization and finetuning, the Generative Diffusion Datasets (GDDs) were used to train the CNN-XGBoost models. The Figure 1. shows the model structure in stage1 and stage2.

**Fig 1:**
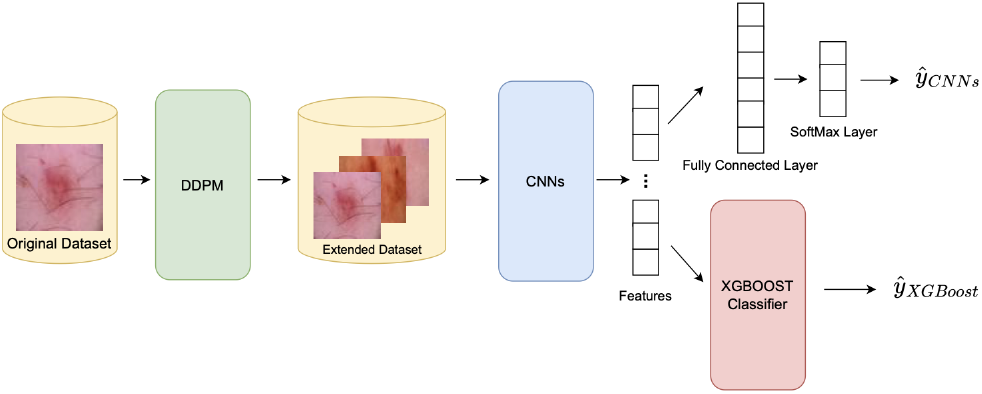
Structure of the DDPM-CNN and DDPM-CNN-XGBoost models. In Stage 1, the model structure comprises the Denoising Diffusion Probabilistic Model (DDPM) and CNN architectures. In Stage 2, the model structure integrates the DDPM, CNN architectures, and the XGBoost classifier.

## III. Results

### A. Dataset characteristics

The original melanoma dataset from Kaggle contains 9,600 images for model training and 1,000 images for evaluation14. Specifically, the training set comprises 4,800 benign and 4,800 malignant melanoma images. Using the generative diffusion model, we constructed multiple Generative Diffusion Datasets (GDDs) based on this training set. Table 1. provides detailed information about these GDDs. The value of lambda (*λ*) represents the relative size of each Generative Diffusion Dataset (GDD), defined as the ratio of synthetic images in the Generative Diffusion Dataset (GDD) dataset to the original training images. When the value of lambda (*λ*) is equal to 0, it means there is not synthetic images in that GDD.

**TABLE I:** Characteristics of the Generative Diffusion Datasets (GDDs)

| $\lambda$ | Malignant (Train) | Benign (Train) | Test | Total |
| --- | --- | --- | --- | --- |
| 0 (Original) | 4,800 | 4,800 | 1,000 | 10,600 |
| 1 | 9,600 | 9,600 | 1,000 | 20,200 |
| 2 | 14,400 | 14,400 | 1,000 | 29,800 |
| 3 | 19,200 | 19,200 | 1,000 | 39,400 |
| 4 | 24,000 | 24,000 | 1,000 | 49,000 |
| 5 | 28,800 | 28,800 | 1,000 | 58,600 |
| 6 | 33,600 | 33,600 | 1,000 | 68,200 |
| 7 | 38,400 | 38,400 | 1,000 | 77,800 |
| 8 | 43,200 | 43,200 | 1,000 | 87,400 |

### B. Stage 1 Performance of using the GDD dataset and different deep learning CNN models

The average accuracy, AUC, and F1 score of the four CNN architectures (ResNet18, ResNet50, VGG11, VGG16) trained without Generative Diffusion Datasets (GDD) for diagnosing melanoma as benign or malignant were 91.1The datasets augmented by GDD significantly improved the melanoma classification performance, as illustrated in Figure 3 (A, B, C) and detailed in Table 2. Specifically, as the value of *λ* increased, the accuracy for ResNet18, ResNet50, VGG11, and VGG16 first rose and subsequently declined. Accuracy for ResNet (18 and 50 layers) models peaked at *λ* = 4, while the VGG (11 and 16 layers) models reached their highest accuracy at *λ* = 2. This indicates that different CNN architectures achieve maximum accuracy at distinct optimal *λ* values. The optimal *λ* values can be found by using early stopping algorithm if there is no enough computing resources. Notably, using GDD datasets at virtually any *λ* value consistently outperformed the original dataset (*λ* = 0), underscoring the effectiveness of DDPM-generated synthetic images. According to Table 2, the best-performing models were ResNet18 (*λ* = 4) and ResNet50 (*λ* = 4), achieving accuracies of 92.8% and 92.9%, respectively. Compared to the baseline average accuracy without GDD datasets, these represent improvements of nearly 2%. Furthermore, compared to baseline values, the ResNet18 (*λ* = 4) model improved AUC by 0.7% and F1 score by 2%, whereas ResNet50 (*λ* = 4) improved AUC by 1.5% and F1 score by 1.7%. The stage 1 results thus clearly demonstrate that DDPM-generated synthetic images significantly enhance CNN model performance for melanoma classification.

**Fig 2:**
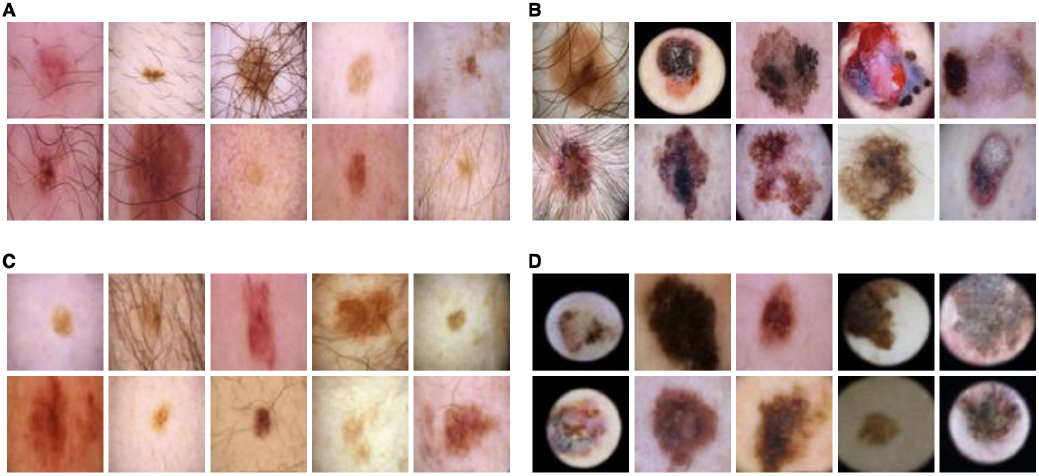
Example images from the original training set and images generated by the Denoising Diffusion Probabilistic Model (DDPM). (A) Benign melanocyte images from the original dataset. (B) Malignant melanocyte images from the original dataset. (C) Synthetic benign melanocyte images generated by DDPM. (D) Synthetic malignant melanocyte images generated by DDPM.

**Fig 3:**
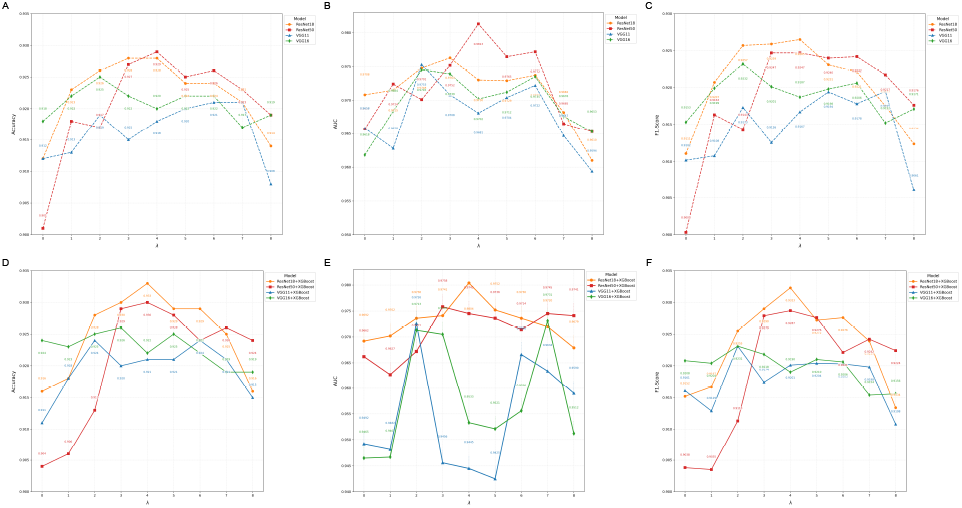
Performance comparison between DDPM-CNN models and DDPM-CNN-XGBoost models. Panels (A–C) show accuracy, AUC, and F1 score for DDPM-CNN models across various *λ* values, while panels (D–F) illustrate these metrics for DDPM-CNN-XGBoost models. Among the DDPM-CNN models, ResNet18 (*λ* = 4) and ResNet50 (*λ* = 4) achieved the highest performance. Overall, the ResNet18+XGBoost model (*λ* = 4) demonstrated the best classification performance.

### C. Stage 2 Performance DDPM-CNN-XGBoost Models

In Stage 1, we used Generative Diffusion Datasets (GDD) to improve melanoma classification accuracy, as shown in Table 1. Classic deep learning models such as ResNet18, ResNet50, VGG11, and VGG16 performed well in this task. In Stage 2, we enhanced the architecture of these four neural network models by replacing their fully connected layers with XGBoost classifiers, creating the CNN-XGBoost models. Stage 2 built on the approach from Stage 1 by using GDD to further improve classification accuracy. Figure 3 (D–F) and Table 3 present the accuracy, AUC, and F1 scores of the CNN-XGBoost models in the melanoma classification task. The CNN-XGBoost approach introduced the XGBoost classifier to further enhance performance. In terms of accuracy, the average accuracy of ResNet18 (*λ*=0), ResNet50 (*λ*=0), VGG11 (*λ*=0), and VGG16 (*λ*=0) with XGBoost was 91.4%, representing a 0.3% improvement compared to the same models without XGBoost. This demonstrates that adding the XGBoost classifier can improve melanoma classification accuracy. As shown in Table 3, the highest accuracies achieved by ResNet18, ResNet50, VGG11, and VGG16 with XGBoost were 93.3%, 93.0%, 92.4%, and 92.6%, respectively. Therefore, ResNet18-XGBoost (*λ*=4) was the best-performing configuration among all DDPM-CNN-XGBoost models and across all experiments. The accuracy of DDPM-ResNet18-XGBoost (*λ*=4) reached 93.3%, reflecting a 2.4% improvement over the average accuracy of the four CNN architectures without GDD and a 0.43% increase over the best model in stage 1. These results in stage2 demonstrate that incorporating the XGBoost classifier significantly improves the system’s performance in melanoma classification.

## IV. DISCUSSION

Skin tumors such as melanoma are highly curable when detected early; however, current diagnostic modalities are not effective for screening large populations to identify suspicious lesions [23]. The advancement of deep learning has significantly accelerated the development of non-invasive diagnostic tools for skin tumors [24]. Recently, several studies have constructed multimodal models for the diagnosis and staging of skin tumors [25] [26] [27].

In this study, we aimed to evaluate the performance of deep learning systems in distinguishing melanoma from benign moles based on a large dataset of labeled tumor photos. The results demonstrated that deep learning algorithms performed well in melanoma classification. The synthetic datasets generated by the Denoising Diffusion Probabilistic Model (DDPM) and the addition of an XGBoost classifier to the classic CNN architectures further improved model performance.

In stage 1, we constructed generative diffusion datasets (GDDs) generated by using DDPM, which contributed significantly to improving classification accuracy. In Stage 2, we replaced the final fully connected layers of the four CNN models with XGBoost classifiers. Loading pre-trained models and applying fine-tuning accelerated convergence and enhanced performance [28]. The best-performing configuration, DDPM-ResNet18+XGBoost with *λ*=4, achieved an accuracy of 93.3%. These findings confirm that our approach is effective in classifying melanoma and has potential for supporting early detection during post-consultation screening.

This work is consistent with Safa Riyadh Waheed’s report, which demonstrated that CNNs can distinguish malignant from non-malignant melanoma after training on large datasets [29]. Both studies showed that classic neural network models achieve high accuracy when trained with substantial data. Notably, the accuracy in our study (93.3%) exceeded that reported by Safa Riyadh Waheed (91%). This improvement is primarily attributed to our use of synthetic data generated by DDPM, which enhanced performance by generating tens of thousands of proportionally balanced training images, and the addition of XGBoost classifiers.

Khalil Aljohani evaluated various modern neural network models for melanoma classification [30]. Like his work, our study used GPU acceleration and large datasets for training. However, we further extended this approach by introducing GDD-CNN-XGBoost models, which achieved an accuracy of 93.3% on the test set (1,000 images), substantially outperforming the 74.9% reported in Aljohani’s work. Of course, the difference may come from the use of different data sets. Khalil Aljohani’s research used sub-class data from multiple authoritative data sets such as ISIC 2017 [31] [32].

Despite promising results, this study has limitations. First, the Kaggle dataset differs from the medical-grade images used in studies such as Zhongwen Li’s or the clinical images reported by Shunichi Jinnai [17] [33]. Those datasets more closely represent real-world diagnostic conditions. Access to such labeled datasets would help further validate and generalize our models. Additionally, our dataset was relatively limited in diversity. Manu Goyal’s work integrated up to 13 melanoma datasets, improving generalizability, which is more universal [34].

Second, although we applied fine-tuning as part of transfer learning, our approach involved training four classic models from scratch on the Kaggle dataset and fine-tuning them afterward. In future work, it may be preferable to adopt pretrained deep models, such as those developed by Andre Esteva, and then fine-tune them for melanoma classification [26]. Their dataset contains 129,450 medical images, in which melanoma is classified in detail. Moreover, evolving methodologies like Explainable Artificial Intelligence (XAI) are enhancing deep learning models through multidimensional optimization, offering valuable insights for future reseach [25].

This study achieved significant results in melanoma classification. Going forward, we plan to explore additional deep learning methods, including integrating linear discriminant analysis (LDA) for feature extraction as demonstrated by Adeniyi, followed by classification using algorithms like XGBoost [35]. This approach could improve interpretability compared to the “black box” nature of neural networks. Additionally, classic transfer learning models such as GoogleNet, MobileNet-v2, and Eatedal Alabdulkreem’s novel lightweight convolutional neural networks (LWCNN) could be applied to this problem [36] [37] [38]. Lightweight networks can save resources and power. If the classification effect on LWCNN is good, it will help to promote it to practical medical applications.

In summary, this study proposed combining generative models with classifier replacement strategies to address the challenge of limited melanoma imaging data. This approach significantly improved test set accuracy (from 91.1% to 93.3%). With highly accurate deep learning models, the early detection rate of malignant skin tumors can be substantially increased, offering important benefits for patient outcomes.

## Data Availability

All data produced in the present study are available upon reasonable request to the authors

## Funding Statement

This work was supported by the Wenzhou-Kean University and Kean University President’s Global-Local (Glocal) Anchor Institution Research Grant Program under ICRPGL2025001.

## Author Contributions

G.T., Y.W., B.Z., and Y.Yu. conceived and designed the experiments. G.T., Y.W., B.Z., and J.L. collected and analyzed the data and produced the experimental results. All authors contributed to the writing and revision of the manuscript (G.T., Y.W., B.Z., Y.Yan, J.L., Yue.W., S.S.K., Q.Y., and Y.Yu.).

## Data Availability Statement

The datasets used for training and evaluation are publicly available on Kaggle at https://www.kaggle.com/dsv/3376422 (DOI: 10.34740/KAGGLE/DSV/3376422).

## Ethics Approval and Consent to Participate

Not applicable.

## Consent for Publication

Not applicable.

## Competing Interests

The authors declare that they have no competing interests.

## Appendix A Side-by-Side Comparison of DDPM-CNN and DDPM-CNN-XGBoost

**TABLE II:** Side-by-side performance comparison of DDPM-CNN and DDPM-CNN-XGBoost models under different *λ* values.

| (a) DDPM-CNN |  |  |  | (b) DDPM-CNN-XGBoost |  |  |  |
| --- | --- | --- | --- | --- | --- | --- | --- |
| Model | Acc. | AUC | F1 | Model | Acc. | AUC | F1 |
| ResNet18 ($\lambda=0$) | 0.912 | 0.9708 | 0.9111 | ResNet18+XGB ($\lambda=0$) | 0.916 | 0.9692 | 0.9152 |
| ResNet18 ($\lambda=1$) | 0.923 | 0.9715 | 0.9207 | ResNet18+XGB ($\lambda=1$) | 0.918 | 0.9702 | 0.9167 |
| ResNet18 ($\lambda=2$) | 0.926 | 0.9748 | 0.9257 | ResNet18+XGB ($\lambda=2$) | 0.928 | 0.9736 | 0.9255 |
| ResNet18 ($\lambda=3$) | 0.928 | 0.9763 | 0.9259 | ResNet18+XGB ($\lambda=3$) | 0.930 | 0.9741 | 0.9290 |
| ResNet18 ($\lambda=4$) | **0.928** | **0.9730** | **0.9265** | ResNet18+XGB ($\lambda=4$) | **0.933** | **0.9804** | **0.9323** |
| ResNet18 ($\lambda=5$) | 0.924 | 0.9729 | 0.9231 | ResNet18+XGB ($\lambda=5$) | 0.929 | 0.9752 | 0.9272 |
| ResNet18 ($\lambda=6$) | 0.924 | 0.9737 | 0.9221 | ResNet18+XGB ($\lambda=6$) | 0.929 | 0.9736 | 0.9276 |
| ResNet18 ($\lambda=7$) | 0.921 | 0.9682 | 0.9175 | ResNet18+XGB ($\lambda=7$) | 0.925 | 0.9720 | 0.9239 |
| ResNet18 ($\lambda=8$) | 0.914 | 0.9610 | 0.9124 | ResNet18+XGB ($\lambda=8$) | 0.916 | 0.9679 | 0.9134 |
| ResNet50 ($\lambda=0$) | 0.901 | 0.9657 | 0.9003 | ResNet50+XGB ($\lambda=0$) | 0.904 | 0.9662 | 0.9038 |
| ResNet50 ($\lambda=1$) | 0.918 | 0.9724 | 0.9163 | ResNet50+XGB ($\lambda=1$) | 0.906 | 0.9627 | 0.9035 |
| ResNet50 ($\lambda=2$) | 0.917 | 0.9701 | 0.9143 | ResNet50+XGB ($\lambda=2$) | 0.913 | 0.9672 | 0.9113 |
| ResNet50 ($\lambda=3$) | 0.927 | 0.9752 | 0.9247 | ResNet50+XGB ($\lambda=3$) | 0.929 | 0.9758 | 0.9279 |
| ResNet50 ($\lambda=4$) | **0.929** | **0.9813** | **0.9247** | ResNet50+XGB ($\lambda=4$) | **0.930** | **0.9745** | **0.9287** |
| ResNet50 ($\lambda=5$) | 0.925 | 0.9764 | 0.9240 | ResNet50+XGB ($\lambda=5$) | 0.928 | 0.9736 | 0.9276 |
| ResNet50 ($\lambda=6$) | 0.926 | 0.9772 | 0.9242 | ResNet50+XGB ($\lambda=6$) | 0.924 | 0.9714 | 0.9221 |
| ResNet50 ($\lambda=7$) | 0.923 | 0.9665 | 0.9217 | ResNet50+XGB ($\lambda=7$) | 0.926 | 0.9745 | 0.9242 |
| ResNet50 ($\lambda=8$) | 0.919 | 0.9653 | 0.9176 | ResNet50+XGB ($\lambda=8$) | 0.924 | 0.9741 | 0.9224 |
| VGG11 ($\lambda=0$) | 0.912 | 0.9658 | 0.9102 | VGG11+XGB ($\lambda=0$) | 0.911 | 0.9492 | 0.9161 |
| VGG11 ($\lambda=1$) | 0.913 | 0.9628 | 0.9108 | VGG11+XGB ($\lambda=1$) | 0.918 | 0.9482 | 0.9129 |
| VGG11 ($\lambda=2$) | 0.919 | 0.9753 | 0.9173 | VGG11+XGB ($\lambda=2$) | **0.924** | **0.9726** | **0.9231** |
| VGG11 ($\lambda=3$) | 0.915 | 0.9708 | 0.9126 | VGG11+XGB ($\lambda=3$) | 0.922 | 0.9456 | 0.9174 |
| VGG11 ($\lambda=4$) | 0.918 | 0.9681 | 0.9167 | VGG11+XGB ($\lambda=4$) | 0.921 | 0.9445 | 0.9201 |
| VGG11 ($\lambda=5$) | 0.920 | 0.9704 | 0.9194 | VGG11+XGB ($\lambda=5$) | 0.921 | 0.9425 | 0.9204 |
| VGG11 ($\lambda=6$) | **0.921** | **0.9722** | **0.9178** | VGG11+XGB ($\lambda=6$) | 0.924 | 0.9666 | 0.9203 |
| VGG11 ($\lambda=7$) | 0.921 | 0.9647 | 0.9195 | VGG11+XGB ($\lambda=7$) | 0.921 | 0.9634 | 0.9198 |
| VGG11 ($\lambda=8$) | 0.908 | 0.9594 | 0.9061 | VGG11+XGB ($\lambda=8$) | 0.915 | 0.9590 | 0.9108 |
| VGG16 ($\lambda=0$) | 0.918 | 0.9618 | 0.9153 | VGG16+XGB ($\lambda=0$) | 0.924 | 0.9465 | 0.9208 |
| VGG16 ($\lambda=1$) | 0.922 | 0.9684 | 0.9199 | VGG16+XGB ($\lambda=1$) | 0.923 | 0.9467 | 0.9204 |
| VGG16 ($\lambda=2$) | 0.925 | 0.9745 | 0.9232 | VGG16+XGB ($\lambda=2$) | 0.925 | 0.9713 | 0.9231 |
| VGG16 ($\lambda=3$) | 0.922 | 0.9739 | 0.9201 | VGG16+XGB ($\lambda=3$) | **0.926** | **0.9705** | **0.9218** |
| VGG16 ($\lambda=4$) | 0.920 | 0.9702 | 0.9187 | VGG16+XGB ($\lambda=4$) | 0.922 | 0.9533 | 0.9190 |
| VGG16 ($\lambda=5$) | 0.922 | 0.9712 | 0.9198 | VGG16+XGB ($\lambda=5$) | 0.925 | 0.9521 | 0.9210 |
| VGG16 ($\lambda=6$) | **0.922** | **0.9735** | **0.9206** | VGG16+XGB ($\lambda=6$) | 0.922 | 0.9556 | 0.9206 |
| VGG16 ($\lambda=7$) | 0.917 | 0.9676 | 0.9152 | VGG16+XGB ($\lambda=7$) | 0.919 | 0.9731 | 0.9154 |
| VGG16 ($\lambda=8$) | 0.919 | 0.9653 | 0.9171 | VGG16+XGB ($\lambda=8$) | 0.919 | 0.9512 | 0.9156 |

## Notes

### Competing Interest Statement

The authors have declared no competing interest.

### Funding Statement

This study was funded by the Wenzhou-Kean University and Kean University President Global-Local (Glocal) Anchor Institution Research Grant Program under ICRPGL2025001.

### Author Declarations

Kaggle at https://www.kaggle.com/dsv/3376422 (DOI: 10.34740/KAGGLE/DSV/3376422).

